# The potential of digital health technologies in African context, Ethiopia

**DOI:** 10.1101/2021.03.27.21254466

**Authors:** Tsegahun Manyazewal, Yimtubezinash Woldeamanuel, Henry M. Blumberg, Abebaw Fekadu, Vincent C. Marconi

## Abstract

The World Health Organization (WHO) recently put forth a Global Strategy on Digital Health 2020 - 2025 with several countries having already achieved key milestones. We aimed to understand whether and how digital health technologies (DHTs) are absorbed in Africa, tracking Ethiopia as a key node. We conducted a systematic review, searching PubMed-MEDLINE, ScienceDirect, African Journals Online, ClinicalTrials.gov, and the WHO International Clinical Trials Registry Platform databases from inception to 02 February 2021 for studies of any design that investigated the potential of DHTs in clinical or public health practices in Ethiopia. This review was designed to inform our ongoing DHT-enabled randomized controlled trial (RCT) (ClinicalTrials.gov ID: NCT04216420). We found 23,897 potentially-relevant citations, among which 47 studies met the inclusion criteria, comprising a total of 594,999 patients, healthy individuals, and healthcare professionals. The studies involved seven DHTs: mHealth (25 studies, 573,623 participants); electronic health records (13 studies, 4,534 participants); telemedicine (3 studies, 445 participants); cloud-based application (2 studies, 2,382 participants); genomics data (1 study, 47 participants); information communication technology (2 studies, 551 participants), and artificial intelligence (1 study, 13,417 participants). The studies targeted six health conditions: maternal and child health (15), infectious diseases (11), non-communicable diseases (3), dermatitis (1), surgery (3), and general health conditions (14). The outcomes of interest were feasibility, usability, willingness or readiness, effectiveness, quality improvement, and knowledge or attitude towards DHTs. Four studies involved RCTs. The analysis showed that although DHTs are a relatively recent phenomenon in Ethiopia, their potential harnessing clinical and public health practices are highly visible. Their adoption and implementation in full capacity requires more training, access to better devices such as smartphones, and infrastructure. DHTs hold much promise tackling major clinical and public health backlogs and strengthening the healthcare ecosystem in Africa. More RCTs are needed on emerging DHTs including artificial intelligence, big data, cloud, genomics data, cybersecurity, telemedicine, and wearable devices to provide robust evidence of their potential use in such settings and to materialize the WHO’s Global Digital Health Strategy.

## INTRODUCTION

Health technology innovations are transforming the discovery, development, and delivery of health products and services^1–4^ and significantly changing the way health conditions are diagnosed, treated and prevented.^5,6^ These innovations are building a sustainable foundation for affordable, accessible, and high-quality medicines, vaccines, medical devices, and system innovations, pursuing novel solutions, entrepreneurial ventures, and public sector efforts to the most challenging health problems.^7,8^ Digital health technologies (DHTs) such as smartphone apps, web-based platforms, and wearable devices are rapidly emerging as promising interventions for disease management,^9–12^ and telemedicine is making remote healthcare more feasible.^13–17^ Artificial intelligence (AI) has the potential to perform many healthcare tasks well (e.g. outcome prediction)^18,19^ and to support the learning and professional development of healthcare professionals.^20,21^ Pharmacogenomics are driving precision medicine into the future^22,23^ and process innovations are improving healthcare delivery through novel approaches to research and clinical trials.^24–27^ The healthcare sector, in general, is one of the most important financiers in innovation, second to the information technology sector. Medical technology patents grew the fastest at close to 6% per year, and pharmaceutical, biotech, and medical device firms are among the top global corporate investors in research and development, spending over 100 billion US dollars annually.^28^ More innovations are expected to emerge as healthcare spending rises, with a global estimated average spending of $11,674 per person by 2022, for reasons including increased costs for healthcare providers, aging population, healthcare coverage, and new treatments and technologies.^29^

Nevertheless, many of these breakthroughs have not reached the healthcare providers and the people most in need to tackle the rising burden of diseases.^30–32^ People living in low-income countries, such as many countries in Africa, are at high risk of many health conditions compared to those living in other regions, while having the most limited access to health innovations.^30,33^ Africa has the greatest healthcare challenges in the world: life expectancy is 60 years, substantially lower than the global average of 72 years; maternal mortality ratio is 547 per 100,000, but 13 in high-income countries and 216 globally; under 5 mortality is 76 per 1,000, but 5 in high-income countries and 39 globally.^34,35^ While there were 1,098 researchers per million inhabitants globally, the corresponding figure for Africa was 87.9 per 1 million. Africa lags in the capacities for health technology innovations, while it bears 23% of the global disease burden and 16% of the world population, with the continent expected to double its population by 2050, from 1 billion to nearly 2.4 billion.^36–40^

Without urgent technological, industrial, intellectual, and research-oriented health interventions, Africa cannot tackle the needs and demands of its population. If health technology innovations are needed to transform health system gaps in Africa, it is important to generate country-specific evidence to identify challenges and opportunities in the region as potential resources for further interventions. The World Health Organization (WHO) embraced a more proactive stance in this regard. In 2020, the WHO developed a global strategy on digital health for 2020-2025.^41^ The vision of the global strategy was to improve health for everyone, everywhere by accelerating the development and adoption of appropriate, accessible, affordable, scalable, and sustainable person-centric digital health solutions to prevent, detect and respond to epidemics and pandemics, developing infrastructure and applications that enable countries to use data to promote health and wellbeing, and to achieve the health-related United Nations’s Sustainable Development Goals (SDGs). Through its Africa office, the WHO Regional Office for Africa (WHO-AFRO), the WHO designed the health technologies and Innovations program to guide the assessments, development, ethics, use, and monitoring of national health technology strategies, with a broader aim of improving access, quality, and rational use of health innovations, including medicines, medical products, and technologies.^42^ Similarly, in 2019, the WHO developed a guideline that established recommendations on DHTs for health systems.

This study is in support of the WHO’s DHT initiatives. We focused on Ethiopia, the fastest growing economy in Africa per the 2019 World Bank report^43^, and the second most populated country, with more than 112 million people in 2019. Ethiopia aims to reach lower-middle-income status by 2025, with strong commitment and dedication to achieve the SDGs by 2030. The Ethiopian Ministry of Health (MOH) recently, on 06 August 2020, launched a Digital Health Innovation and Learning Center, the first of its kind, where experts can design and validate digital health tools, synthesize and promote best practices, and scale-up innovations.^44^ Evidence on the services and application potential of DHTs has not been synthesized in the country to inform debates and decisions.

Thus, we aimed to investigate whether and how digital health technologies (DHTs) are absorbed in Africa, tracking Ethiopia as a key node.

## METHODS

### Study design

This analysis was designed to provide an initial synthesis of a DHT-enabled randomized controlled trial (RCT) on tuberculosis being conducted in Ethiopia (ClinicalTrials.gov, ID: NCT04216420). This study was based on a systematic review of scientific literature utilizing the Preferred Reporting Items for Systematic Review and Meta-Analysis Protocols (PRISMA-P) 2015 guidelines for the design and reporting of the results (see checklist in the supplementary additional file 1).

To broaden the scope of DHTs in our review, we combined the latest descriptions of digital health given by the WHO^41^ and the U.S. Drug and Food Administration (FDA).^45^ With this, the following technologies were included in the review: mobile health, telehealth, electronic health records (EHR), telemedicine, health information technology, wearable devices, software as a medical device, artificial intelligence, machine learning, genomics, computing sciences in big data, cybersecurity, wireless medical devices, and medical device interoperability.

### Search strategy

We searched the PubMed-MEDLINE, ScienceDirect, African Journals Online, ClinicalTrials.gov, and the WHO International Clinical Trials Registry Platform databases from inception to the latest 02 February 2021 for studies of any design and in any setting in Ethiopia that investigated the potential of DHTs in clinical or public health practices in Ethiopia. We performed manual searches of the WHO website, Google search engine, and reference lists of included studies, and contacted authors of original studies to retrieve extra possible articles or additional data (see search strategy in the supplementary additional file 2).

We tailored search strategies to each database and used controlled medical subject headings (MeSHs) and search filters where available, or Boolean search methods and free-text terms, referring to Ethiopia and Digital OR Mobile OR Smartphone OR “Cell phone” OR Techno* OR “short message service” OR SMS OR Tele* OR Telemedicine OR Telehealth OR E-health OR eHealth OR Remote OR Electro* OR Comput* OR cloud OR Software OR Application OR Robotics OR Blockchain OR “Artificial intelligence” OR genomics OR “big data” OR cybersecurity OR wireless.

### Eligibility

Studies were included if they met the following inclusion criteria:

#### Participants

Eligible participants could be patients, healthcare professionals, data collectors, or healthy individuals in Ethiopia, either women or men, and without age restrictions. Thus, the search was not restricted to participants except that they should reside in Ethiopia.

#### Interventions

All DHTs that were included in our definition. Studies that investigated digital technologies for non-health conditions were excluded.

#### Comparisons

Studies with a comparison condition were not required as a criterion. Thus, studies with or without a comparator were eligible.

#### Outcome

Studies assessing the potential efficacy, effectiveness, feasibility, usability, acceptability, or any related outcomes were included in the review without specific restrictions.

#### Study design

All available study designs were included. We excluded reviews, commentaries, editorials, and proceedings as these are non-empirical publications.

## Study selection

Two independent authors examined the title and abstract of all screened publications. From the title and abstract of all publications identified by the database search, those that were duplicated or did not meet the inclusion criteria were excluded. The full texts of the remaining publications were further reviewed. Disagreements were resolved by consensus and, if persisted, were arbitrated through discussion with a third author.

### Data extraction

The identified data were listed, and information was provided on the type and details of the type of DHT under investigation, the disease condition studied, the type of participants, the number of participants, the study design employed, outcome measures, major findings, the surname of the first author, and year of publication.

### Data management and analysis

The publications were grouped in exhaustive tables based on the type of DHT investigated. A qualitative content analysis of all documents and articles was performed. Each article was summarized, and the data were reported descriptively. Less emphasis was placed on the assessment of the quality of the included literature as that was not the major objective of this review.

### Operational definitions

Mobile health (mHealth): The use of mobile phone device’s core utility of voice or short messaging service (SMS) as well as more complex functionalities to improve health outcomes and healthcare services.

Electronic health records (EHRs): are patient-centered electronic records that provide immediate and secure information to authorized users.

Telemedicine: The practice of medicine at a distance which involves an interaction between a healthcare provider and a patient when the two are separated by distance.

Cloud: The practice of storing and computing health data remotely over the internet, which is managed by external service providers.

Genomic database: An online repository of genomic variants for storing, sharing and comparison of data as a medical and scientific resource.

Artificial intelligence (AI): The simulation of human intelligence in a digital computer that is programmed to think or perform health tasks like humans.

Information and communications technology (ICT): technologies that provide access to health information through telecommunications

## RESULTS

### Characteristics of included studies

#### Study selection

From the 23,897 articles screened, 23,549 were excluded based on the title or abstract. The rest of 203 full-text articles were screened for eligibility, of which 156 were excluded for being irrelevant to the main subject (122) or focused on non-health conditions (34). Forty-seven studies were identified that met the inclusion criteria. **Figure 1** summarizes the PRISMA flowchart of the study.

**Figure 1:**
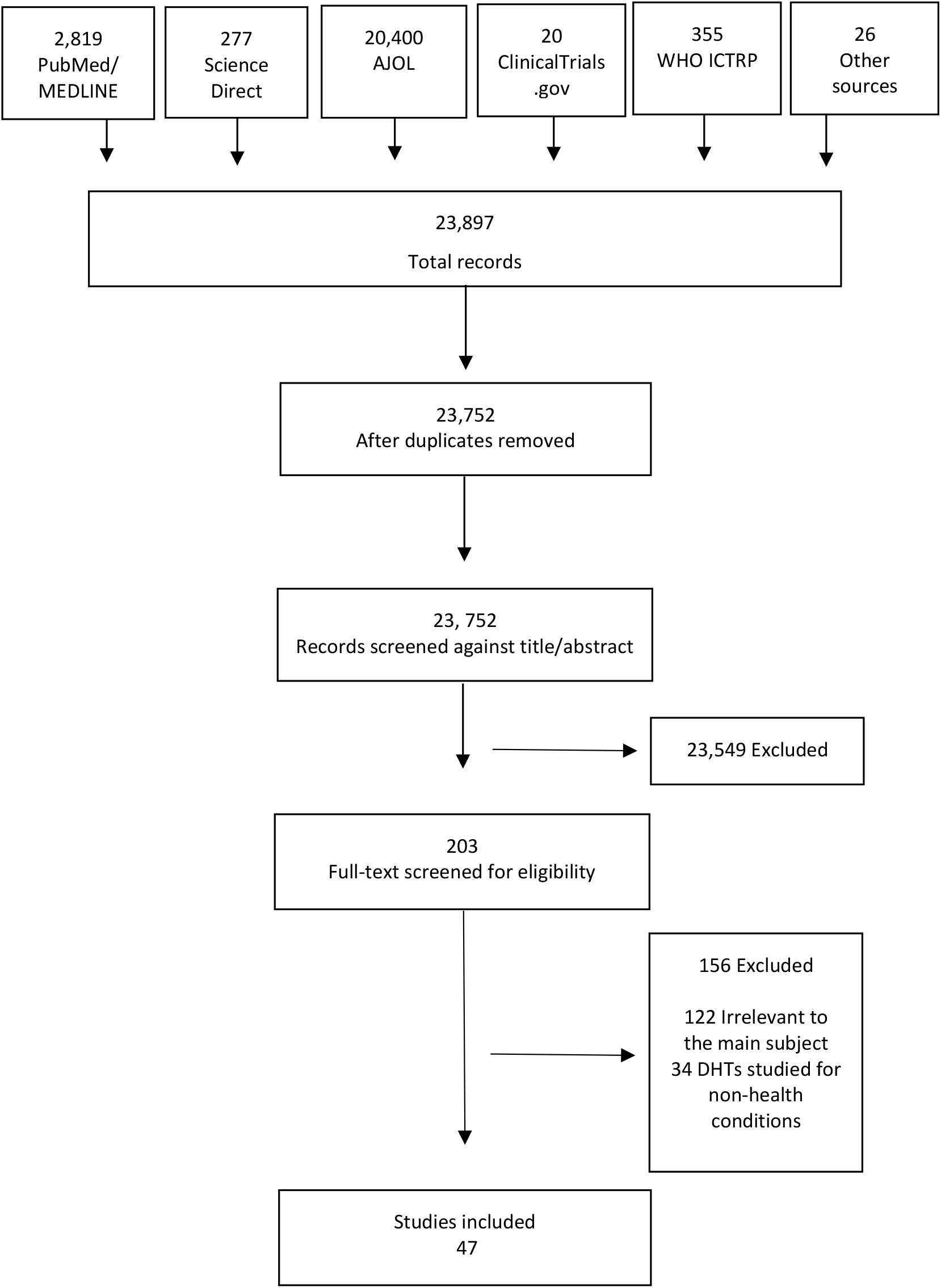
PRISMA flow diagram of the study

#### Participants

The 47 included studies had a total of 594,999 study participants. The health conditions studied were maternal and child health, including antenatal care, postnatal care, infant feeding, contraceptives, and delivery (n = 15), infectious diseases including TB, HIV, malaria, lymphatic filariasis, and onchocerciasis (n = 11), non-communicable diseases including diabetes and cancer (n = 3), dermatitis (n = 1), surgery (n = 3), and the remaining 14 had general health conditions.

#### Interventions

The included studies were involved seven digital health domains: 25 on mobile health (mHealth) (573,623 participants); 13 on EHR (4,534 participants); three on telemedicine (445 participants); two on Cloud usage (2,382 participants); one on genomics (47 participants), two on information and communication technology (ICT, 551 participants), and one on AI (13,417 participants). Our search did not find articles relevant to wearable devices, software as a medical device, computing sciences in big data, cybersecurity, wireless medical devices, and robotics.

#### Outcomes

The primary outcome variables were feasibility (n = 9), usability (n=9), willingness or readiness to use (n = 10), effectiveness (n = 8), quality improvement (n = 7), and knowledge or attitude about the DHT (n = 4). Some of the studies employed two or more of these outcomes.

#### Study type

The study designs were cross-sectional study (n = 37), RCT or non-randomized experimental (5), and cohort (5).

#### Study setting

All the studies were conducted in Ethiopia. Some of the studies were multi-country with a significant number of participants included from Ethiopia.

## DIGITAL HEALTH TECHNOLOGIES

### mHealth

There were 25 publications identified in the mHealth field,^46–70^ involving a total of 573,623 study participants: 401,563 patients, 171,295 healthy individuals, and 765 healthcare professionals. The most common citations were on the potential use of mHealth for maternal, child, and reproductive health, covering 60% (15/25) of the citations, followed by infectious diseases, 44% (11/25). The publications were mainly cross-sectional, 68% (17/25), followed by RCT, 16% (4/25). The outcomes of interest were effectiveness (n = 7), usability (n = 5), feasibility (n = 4), quality (n = 4), willingness (n = 4), and knowledge (n = 1). **Table 1** summarizes the characteristics of the included mHealth studies.

**Table 1:** Characteristics of included mHealth studies (n = 25)

| Reference | mHealth | Condition | Participants type | Participants # | Study design | Outcome measure | Finding |
| --- | --- | --- | --- | --- | --- | --- | --- |
| Yigezu et al. 2020<sup>46</sup> | Mobile-based VCT | HIV VCT | VCT attendants | 144,267 | Cross-sectional - cost-effectiveness | Effectiveness – cost-effectiveness | Mobile-based VCT costs less than both facility-based and stand-alone VCTs |
| Gebremariam et al 2020<sup>47</sup> | SMS | Infant feeding | Parents of child-bearing age | 41 | Cross-sectional | Feasibility, acceptability | Feasible and acceptable option for knowledge sharing and awareness |
| Starr et al 2020<sup>48</sup> | mHealth | Post-surgery follow-up | Patients on post-surgery follow-up | 701 | Cohort, prospective | Feasibility | Telephone follow-up after surgery is feasible and valuable |
| Jadhav et al 2020<sup>49</sup> | Own mobile phone | Contraceptive | Women of reproductive age | 15,683 | Cross-sectional, retrospective | Effectiveness- Contraceptive uptake | No association between mobile phone ownership and contraceptive uptake |
| Bradley et al 2020<sup>50</sup> | Smartphone | Post-surgery follow-up | Patients on post-surgery follow-up | 24 | Cohort | Feasibility | smartphones were low-cost, reliable method to follow-up patient after surgery |
| Tadesse et al 2019<sup>51</sup> | mHealth-based e-Partograph | Obstetric care | Healthcare professionals | 466 | Cross-sectional | Willingness | 46% willing to use mobile-phone for e-Partograph |
| Kassa et al 2019<sup>52</sup> | Own mobile phone | Postnatal care | Women in postnatal care | 370 | Cross-sectional | Knowledge, attitude | 3x higher odds of positive attitude to preconception in women who own phone |
| Kebede et al. 2019<sup>53</sup> | SMS or voice call reminder | Postnatal appointment | Women in postnatal care | 700 | RCT | Effectiveness - Postnatal compliance | 3x higher odds of postnatal compliance in women who received a reminder |
| Thomsen et al 2019<sup>54</sup> | mHealth-based Safe Delivery App | Delivery | Healthcare professionals | 56 | Cross-sectional | Usability - user experience | The App improved providers' delivery knowledge and skills |
| Jemere et al 2019<sup>55</sup> | mHealth-based health services | Diabetes | Patients with diabetes | 423 | Cross-sectional | Willingness, access, | 78% had a phone; 71% willing to receive mHealth-based diabetes services |
| Endehabtu et al 2018<sup>56</sup> | SMS-based intervention | Antenatal care | Women in antenatal care | 416 | Cross-sectional | Willingness access, | 36% had smartphones; 71% willing to receive SMS-based antenatal care intervention |
| Mengesha et al. 2018<sup>57</sup> | mHealth-based HMIS | Data use | Health extension workers | 62 | Cross-sectional | Data quality, user experience | mHealth-based HMIS improved data quality, data flow, patient follow-up. |
| Steege et al 2018<sup>58</sup> | mHealth-based data and reminder | TB | Health extension workers | 19 | Cross-sectional | Quality-healthcare delivery | Improved community TB and maternal health service delivery |
| Martindale et al 2018<sup>59</sup> | MeasureSMS-morbidity reporting tool | lymphatic filariasis, podoconiosis | Healthcare professionals | 59 | Cross-sectional, comparative | Effectiveness, cost, time | MeasureSMS tool was more effective, 13.7% less costly than paper-based reporting |
| Abate et al 2018<sup>60</sup> | Telepathology Acquiring microscopic images using a smartphone camera | blood cell count, malaria lab diagnosis | Healthcare professionals | 2 | Cross-sectional | Usability, accuracy | It was fast, cost-effective, and accurate in low resource setting. |
| Shiferaw et al 2018<sup>61</sup> | mHealth-based data collection | Maternal health service | Healthcare professionals | 15 | Experimental/ Implementation | Effectiveness | Timely and complete maternal health data |
| Atnafu et al. 2017<sup>62</sup> | SMS-based data exchange Ap. | Antenatal care | Women on antenatal care | 3240 | RCT | Effectiveness - MCH outcomes | 9% increased deliveries attended by skilled health workers |
| Mablesen et al. 2017<sup>63</sup> | MeasureSMS-Morbidity reporting tool | Lymphatic filariasis (LF) case estimate | People with LF clinical manifestations | 400,000 | Cross-sectional | Usability as a reporting tool | The tool improved survey and reporting of clinical burden of LF |
| Medhanyie et al 2017<sup>64</sup> | Smartphones for collecting patient data | Maternal health records | Healthcare professional | 25 | Cross-sectional | Usability | 8% improved data completeness compared with paper records |
| Shiferaw et al 2016<sup>65</sup> | Locally customized mHealth App. | Delivery and postnatal care | Women on ANC | 2,261 | Cohort | Quality - ANC services utilization | The App improved delivery in health centers, but not ANC visits |
| Lund et al 2016<sup>66</sup> | mHealth safe delivery App (SDA) | Perinatal and neonatal survival | Women in active labor, provider | 3777 | RCT | Quality - Perinatal mortality | The SDA nonsignificantly lowered perinatal mortality compared with standard |
| Kebede et al 2015<sup>67</sup> | SMS medication reminders | HIV | HIV patients on ART | 415 | Cross-sectional | Willingness, access | 76% owned cellphone<br>50.9% willing to receive SMS medication reminder |
| Medhaniye et al 2015<sup>68</sup> | Smartphone-based data records | Maternal health | Healthcare professionals | 24 | Cross-sectional | Usability | The records were useful for day-to-day maternal healthcare services delivery |
| Desta et al 2014<sup>69</sup> | mVedio for behavior change | Maternal and newborn health | Community members | 540 | Cross-sectional. | Effectiveness - Community behavior change | mViedo changed community behavior change on maternal and newborn health in rural Ethiopia |
| Little et al 2013<sup>70</sup> | Smartphone open-source health App. | maternal health | Healthcare professionals | 37 | Cohort | Feasibility - Technical needs | Ownership and empowerment are prerequisites for a successful mHealth program |

The studies had emerging insights into the potential use of mHealth to transform healthcare and improve access to services by addressing financial, social, or geographic factors, though the findings were not consistent. In a study that compared the cost-effectiveness of facility-based, stand-alone, and mobile-based HIV voluntary counseling and testing, the results revealed a cost-effective and improved VCT service with the use of mobile phones when compared with the two arms.^46^ A study that assessed the potential of telephone calls to identify and follow-up post-surgery infections and total complications reported the telephone calls were feasible and valuable.^48^ A similar study that assessed the potential of cell phones to follow-up patients after short-term surgical missions found the tool as cost-effective and reliable.^50^ The use of SMS-based education was found to be a good option for improving the knowledge and awareness of parents regarding infant feeding.^47^

Several studies assessed the potential of using mHealth for family planning and maternal and child health services. One study reported that a high proportion of pregnant women in an antenatal care clinic had a mobile phone and were willing to receive an SMS text message-based mHealth intervention, though two-third of the participants lacked smartphones to upload some application software.^56^ Locally customized mHealth applications during antenatal care significantly improved delivery and postnatal care service utilization through positively influencing the behavior of health workers and their clients.^65^ Mobile phone reminders were effective in terms of enhancing adherence to postnatal care appointments, with the potential improving postnatal appointment adherence.^63^ On the contrary, a retrospective analysis of demographic and health survey data reported that mobile phone ownership or receiving family planning information via SMS had no significant effect on improving contraceptive uptake.^49^

A study that assessed the potential of mHealth on antiretroviral (ART) services for people with HIV reported that the willingness of patients to receive SMS in support of medication reminders was not consistent, with 49% unwilling to receive the reminders.^67^ Age, educational status, and previous experience using the internet had a significant association with their willingness.^67^ A cross-sectional study of patients with diabetes showed that a high proportion of the patients had access to mobile phones and were willing to use them for medication reminders.^55^

Nine of the 25 included studies on mHealth had ‘healthcare professionals’ as their target participants. Several of these studies revealed that mHealth had the potential to improve skills and competency of healthcare providers toward safe birth,^54^ data quality and flow,^57^ patient follow-up,^57^ community-based tuberculosis and maternal health service delivery,^58^ quality and cost-effective reporting of lymphatic filariasis, and podoconiosis data,^59^ and timely and complete reporting of maternal health data.^61,64,68,70^ Some healthcare providers were able to appropriately use mHealth technologies for patient assessment and routine data collection with minimal training and supervision. However, there were major preconditions needed for the healthcare providers to effectively apply such technologies, including in-service application training,^51^ strong connectivity and electric power supply, especially in rural areas of the country,^57^ and uninterrupted mobile network airtime.^68^

### Electronic health records

Of the final set of 47 studies, 13 were on EHR,^71–83^ involving 4,534 study participants: 4,232 health professionals, 250 patients, and 52 healthy individuals. The studies used health information technology, health management information system (HMIS), tablet-based electronic data capture (EDC), electronic information source (EIS), health smart card (HSC), and Android-based data collection system as their EHR of interest. Five studies focus on infectious diseases, one on non-communicable diseases (diabetes), and the remaining seven had a broader health system scope. The studies were all cross-sectional, 92% (12/13), except for one RCT. The outcomes of interest were willingness (n = 5), usability (n = 3), quality (n = 3), and feasibility (n = 2). **Table 2** summarizes the characteristics of the included EHR studies.

**Table 2:** Characteristics of included EHR studies (n = 13)

| Reference | EMR | Condition | Participants type | Participants # | Study design | Outcome measure | Finding |
| --- | --- | --- | --- | --- | --- | --- | --- |
| Seboka et al 2021<sup>71</sup> | Information system for managing diabetes | Diabetes | Healthcare professionals | 406 | Cross-sectional | Willingness, attitude, | 64% had a favorable attitude to remotely monitor diabetes patients, 74% willing to use voice calls. |
| Berihun et al 2020<sup>72</sup> | EMR in health facilities | HIV | Healthcare professionals | 616 | Cross-sectional | Willingness | 86% willing to use EMR |
| Ahmed et al. 2020<sup>73</sup> | EMR in health facilities | - | Healthcare professionals | 420 | Cross-sectional | Willingness intention | 40% intention to use EMR |
| Kebede et al 2020<sup>74</sup> | HMIS in health facilities | - | Healthcare professionals | 332 | Cross-sectional | Quality | 48% accuracy and 82% completeness of data; below national standards |
| Awol et al 2020<sup>75</sup> | EMR in health facilities | - | Healthcare professionals | 414 | Cross-sectional | Willingness-readiness | 62% ready to use EMR system |
| Zelege et al 2019<sup>76</sup> | Electronic data capture (EDC)- tablet | - | Interviewers | 12 | RCT | Quality of data | Better data quality and efficiency with EDC than standard paper-based data |
| Abiy et al. 2018<sup>77</sup> | EMR at ART clinic | HIV | Patients on HIV care | 250 | Cross-sectional, comparative | Quality - completeness, reliability | Slightly lower (76%) data completeness in EMR, than paper-based (78%) |
| Bramo et al 2017<sup>78</sup> | Electronic information source (EIS) | HIV/AIDS Care and Treatment | Healthcare professionals | 352 | Cross-sectional | Usability - utilization | 67% not used EIS for not having training, prefer print resource |
| Dusabe-Richards et al 2016<sup>79</sup> | HMIS | TB | Healthcare professionals | 90 | Cross-sectional | Feasibility | HMIS is usable, but with gaps in quality, accuracy, reliability, timeliness of data |
| Samuel et al 2016<sup>80</sup> | Electronic Information Sources (EIS) | - | Healthcare professionals | 590 | Cross-sectional | Usability, access | 42% used EIS, affected by computer literacy, access to internet |
| Tilahun et al. 2015<sup>81</sup> | SmartCard | - | Healthcare professionals | 406 | Cross-sectional | Usability – user satisfaction, | 61% dissatisfied with the EMR; 64% believed EMR had less quality impact |
| Biruk et al 2014<sup>82</sup> | EMR |  | Healthcare professionals | 606 | Cross-sectional | Willingness - readiness | 54% ready to use EMR |
| King et al 2013<sup>83</sup> | Android-based data collection | Neglected tropical diseases | Community members (households) | 40 | cross-sectional, comparative | Feasibility, effectiveness | Suitable, accurate, and save time over standard paper-based survey questionnaires |

According to the studies reviewed, EHRs have the potential to exchange real-time patient-related data for better clinical decision-making and to capture and share electronic health information efficiently. However, the studies also reported potential gaps and drawbacks associated with EHRs. A study that compared EHR with paper-based records for ART data reported a higher incomplete data with the use of EHR for various reasons including difficulties implementing EHR in high patient load conditions and the frequent need for capturing dual electronic and paper-based data for individual patients.^77^

Studies that assessed the current HMIS practice in healthcare facilities reported several gaps in the accuracy, completeness, and timeliness of data for reasons including poor support from facility management, lack of accountability for data errors, poor supportive supervision, and absence of a dedicated Information Management unit responsible for EHR functions.^74,79^

A significant number of healthcare professionals were either not ready^75,82,78^ or not willing^72,81^ to use EHR. Some healthcare professionals still preferred paper-based records to EHR in their daily work^74,77,79,80,81^ while other preferred EHR.^76^ Lack of access to EHR training, computer skills, and performance expectancy were the major barriers to their willingness or intention to use EHR.^71,72,73,74,75,77,78,81,82^

### Telemedicine

Of the 47 studies considered, three reported on telemedicine, involving 445 participants who were healthcare professionals.^84–86^ The studies investigated what level of knowledge and attitude healthcare professionals have toward telemedicine,^84^ why healthcare providers resist using telemedicine,^85^ and how teledermatology system can be improved in a given program.^86^ All were cross-sectional and their outcomes of interest were knowledge and attitude, willingness, and usability (**Table 3**).

**Table 3:** Characteristics of included telemedicine studies (n = 3)

| Reference | Telemedicine | Condition | Participants type | Participants # | Study design | Outcome measure | Finding |
| --- | --- | --- | --- | --- | --- | --- | --- |
| Biruk et al 2018<sup>84</sup> | Telemedicine | - | Healthcare professionals | 312 | Cross-sectional | Knowledge, attitude | 62% lack good knowledge, 36% lack a good attitude towards telemedicine |
| Xue et al 2015<sup>85</sup> | Telemedicine | - | Healthcare professionals | 107 | Cross-sectional | Willingness - reasons for resistance to telemedicine | Reduced autonomy, anxiety, and costs increased resistance |
| Delaigue et al 2014<sup>86</sup> | Teledermatology | Dermatitis | Healthcare professionals | 26 | Cross-sectional | Usability | Teledermatology delivered a useful service, system gap for case followup |

The studies highlighted that telemedicine has the potential improving healthcare; however, healthcare providers had less knowledge and information about it. One study reported that of the 312 healthcare professionals included, 62% lacked good knowledge and 36% lacked a good attitude about telemedicine.^84^ Healthcare professionals resisted the use of telemedicine in their clinical practices mainly due to their perceived threat and controllability, with reduced autonomy, anxiety, and costs indirectly aggravating the resistance.^85^

### Cloud-based applications

Two studies reported on Cloud-based interventions, involving 2,382 participants: 1,748 surgical cases^87^ and 634 healthy women on cervical cancer screening.^88^ The aims of the studies were on the feasibility of a multicentre Cloud-based peri-operative registry for surgical care^87^ and the feasibility of a cloud-based electronic data system for human papillomavirus (HPV) cervical cancer screening.^88^ **Table 4** summarizes the characteristics of the included Cloud studies.

**Table 4:**
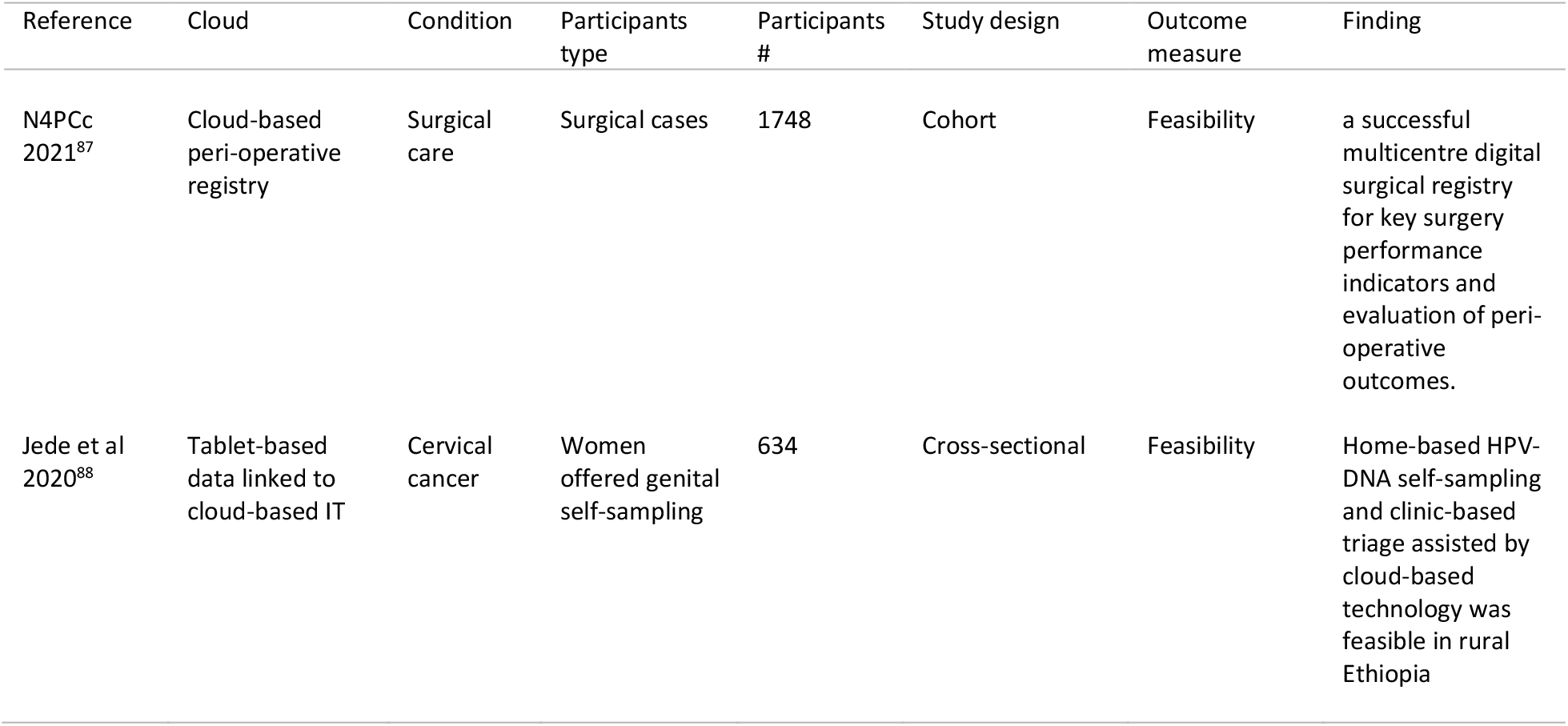
Characteristics of included Cloud-based studies (n = 2)

| Reference | Cloud | Condition | Participants type | Participants # | Study design | Outcome measure | Finding |
| --- | --- | --- | --- | --- | --- | --- | --- |
| N4PCc 2021<sup>87</sup> | Cloud-based peri-operative registry | Surgical care | Surgical cases | 1748 | Cohort | Feasibility | a successful multicentre digital surgical registry for key surgery performance indicators and evaluation of peri-operative outcomes. |
| Jede et al 2020<sup>88</sup> | Tablet-based data linked to cloud-based IT | Cervical cancer | Women offered genital self-sampling | 634 | Cross-sectional | Feasibility | Home-based HPV-DNA self-sampling and clinic-based triage assisted by cloud-based technology was feasible in rural Ethiopia |

A Network for Peri-operative Critical care (N4PCc) developed and evaluated a multicentre Cloud-based peri-operative registry in Ethiopia.^87^ The authors reported on 1748 consecutive surgical cases for key performance indicators including compliance with the World Health Organization’s Surgical Safety Checklist, adverse events during anesthesia, and surgical site infections. With these, the authors reported a successful multicentre digital surgical registry that can enable measurement of key performance indicators for surgery and evaluation of peri-operative outcomes.^87^

One study conducted home-based human papillomavirus (HPV) self-sampling assisted by a Cloud-based electronic data system. The study used an electronic app-based data system with an offline mode function for tablet computers, based on a Cloud solution. The app-based data system showed robust technical functionality, stability, comfort, data accuracy, and ease-of-use by health workers, with no data loss observed. The off-line data collection, uploading, and synchronization system were safe and error-free.^88^

### Genomics data

One DHT study reported on genomics data.^89^ A psychiatric genomics consortium has made an exchange of data between research groups for genome-wide association studies on neurogenetics of schizophrenia, with tens of thousands of patients and controls included. The consortium developed and used the MiGene Family History App (MFHA) to assist clinicians with the collection and analysis of patient genetic data over six months and assessed its feasibility through a survey of 47 clinicians (**Table 5**). The results showed the potential expansion of medical genetics services into low and middle-income countries (LMICs) and the feasibility and benefit of the MFHA for the services.

**Table 5:** Characteristics of included genomics study(n = 1)

| Reference | Genomics | Condition | Participants type | Participants # | Study design | Outcome measure | Finding |
| --- | --- | --- | --- | --- | --- | --- | --- |
| Quinonez et al 2019<sup>89</sup> | MiGene Family History App | Medical genetics services | Healthcare professionals | 47 | Cross-sectional | Feasibility | The App was useful for the collection and analysis of genetics data. |

### Artificial intelligence

One study evaluated the precision of AI in differentiating between target and implanted intraocular lens (IOL) power in cataract outreach campaigns in Ethiopia.^90^ The study applied machine learning (ML) to optimize the IOL inventory and minimize avoidable refractive error in patients from the cataract campaigns (n = 13,417). The result indicated good precision, with the ML optimized the implanted intraocular lens inventory and minimized avoidable refractive error (**Table 6**).

**Table 6:** Characteristics of included artificial intelligence study (n = 1)

| Reference | Artificial intelligence | Condition | Participants type | Participants # | Study design | Outcome measure | Finding |
| --- | --- | --- | --- | --- | --- | --- | --- |
| Brant et al 2020<sup>90</sup> | Machine learning (ML) | Cataract surgery | Cataract patients with target and implanted intraocular lens | 13,417 | Cross-sectional | Effectiveness - precision | ML optimized implanted intraocular lens inventory, minimized avoidable refractive error |

### Information Communication Technology (ICT)

Two studies reported on ICT, involving a total of 551 healthcare professionals (**Table 7**).^91,92^ One study^91^ assessed the digital competency of healthcare providers in seven public health centers and found low-level competency, with factors such as sex, educational status, profession type, monthly income, and years of experience were statistically significant predictors. The second study^92^ assessed health professional’s behavioral intention to adopt eHealth systems and revealed that among the different eHealth constructs, healthcare professionals’ attitude towards eHealth had the strongest effect on the intention to use eHealth systems.

**Table 7:**
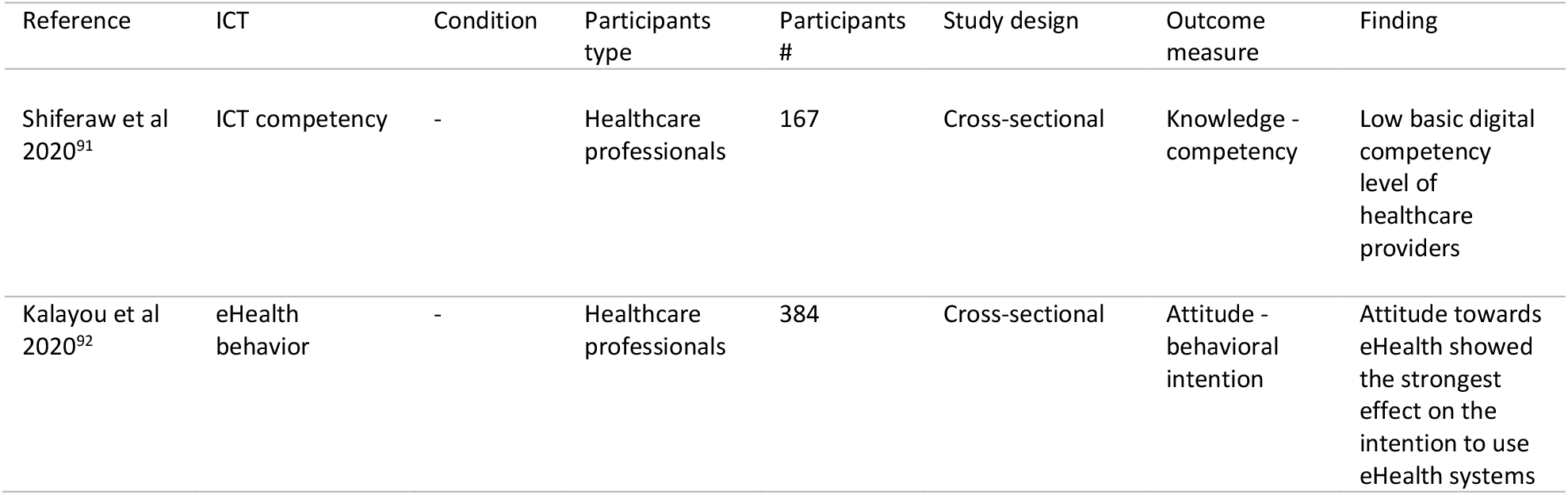
Characteristics of included ICT studies (n = 2)

| Reference | ICT | Condition | Participants type | Participants # | Study design | Outcome measure | Finding |
| --- | --- | --- | --- | --- | --- | --- | --- |
| Shiferaw et al 2020<sup>91</sup> | ICT competency | - | Healthcare professionals | 167 | Cross-sectional | Knowledge - competency | Low basic digital competency level of healthcare providers |
| Kalayou et al 2020<sup>92</sup> | eHealth behavior | - | Healthcare professionals | 384 | Cross-sectional | Attitude - behavioral intention | Attitude towards eHealth showed the strongest effect on the intention to use eHealth systems |

## DISCUSSION

We conducted a systematic review of the available literature to provide strong evidence on the potential impact of DHTs on clinical and public health practices in the context of a resource-constrained sub-Saharan African country, Ethiopia.. The review identified 47 studies across different areas of DHTs, including mHealth, EMR, telemedicine, cloud-based technology, genomic database, ICT, and AI. The analysis showed that though DHTs are a relatively recent phenomenon in Ethiopia, their potential to harness in health systems is highly visible. Digital health solutions have substantial benefits and huge potential to transform the health care system and societal wellbeing in Ethiopia. However, their adoption and implementation in full capacity face challenges in terms of infrastructure, training, access to better devices such as smartphones, and some hesitancies from patients and providers. Such challenges have been reported in studies from other African countries including Uganda,^93,94^ Kenya,^95,96,97,98^, and Tanzania.^99,100^

Of the 47 included studies, emerging DHTs had a very small share at 14%: genomics 2%, AI 2%, Cloud-based technology four percent, and telemedicine six percent, while the major 86% share was for mHealth (53%), EHR (27%), and ICT 6%). This analysis demonstrated that only 9% (4/47) of the studies were tested in RCTs to provide robust and more credible evidence of the potential of the DHTs. A meta-analysis was not conducted due to the heterogeneous nature of the compiled studies.

The mHealth solutions identified in this systematic review mainly aimed to improve maternal and child health care and services. This review found that mHealth interventions, either a phone call or SMS, were feasible and acceptable for improving contraceptive uptake, maternal healthcare, and tuberculosis medication adherence among the Ethiopian population. The evidence also showed evidence of the potential utility of mHealth for HIV counseling and testing, outpatient follow-up, post-surgery follow-up, child-immunization follow-up, pregnant women antenatal and postnatal follow-ups, and in improving knowledge and awareness of parents regarding infant feeding, while its potential for contraceptive uptake was not significant. The included studies revealed that a significant number of study participants owned mobile phones and were willing to participate in mHealth-related clinical or public health interventions. However, the type of mobile phone that the patients own may not be smartphones to support an upload of needed software. Such challenges were also reported elsewhere in Kenya^101,102,103^ and Tanzania^104^ where there exist similar socioeconomic disparities in mobile phone ownership in support of the implementation of mHealth.

Our analysis indicates that patients with HIV may resist having their ART adherence information followed-up using electronic medication reminders for fear of potential disclosure of their HIV status. The finding was consistent with a recent study in Tanzania that fear for potential involuntary disclosure of HIV status significantly affects mHealth interventions in such patients.^105^ The development of the next generation of mHealth in such developing countries requires a broad understanding of the local social contexts that may affect the successes of DHTs.^106^

Our analysis indicates that the use of cloud computing could help resource-constrained countries like Ethiopia to acquire advanced data storage, servers, and databases without investing in new IT infrastructure, though we have identified only two studies that are less likely to support its potential. There have been controversies on the potential benefits of the Cloud and the issues surrounding legal and regulatory implications.^107^ For countries like Ethiopia that have not yet established a standardized legal cybersecurity framework, strategy, and governance at the national level,^108^ adopting appropriate laws and building technical capacity would reassure the partnership and uptake of Cloud services.

Telemedicine was an emerging technology in the Ethiopian health care system which had its drawbacks on successful implementations, despite positive energy that healthcare providers to step up. Building capacity of the healthcare providers before full-scale implementation could bring real benefits out of telemedicine. In the COVID-19 pandemic that restricted physical contacts,^109^ telemedicine revealed significant contributions in Ethiopia by connecting patients with their healthcare providers to discuss and followup their disease conditions.^110^

In recent years, capacities for research, development, and trade on DHTs are rising sharply, while more work is needed to delineate the mechanisms of how the gains could be shared out with resource-constrained countries and global digital health strategy met. Our analysis demonstrated the feasibility and potential demands of DHTs, with the greatest opportunities in emerging health technology markets in Africa.

## CONCLUSION

DHTs hold much promise tackling major clinical and public health backlogs and strengthening health systems in Africa. More RCTs are needed on emerging DHTs including artificial intelligence, big data, cloud, genomics data, cybersecurity, telemedicine, and wearable devices to provide robust evidence of their potential use in such settings and to materialize the Global Digital Health Strategy.

## Data Availability

Data used for all analyses can be found in the Supplementary Files.

## FUNDING

Fogarty International Center and National Institute of Allergy and Infectious Diseases of the US National Institutes of Health (D43TW009127) and the Emory Center for AIDS Research (P30 AI050409). The content is solely the responsibility of the authors and does not necessarily represent the official views of the National Institutes of Health or the Emory Center for AIDS Research.

## AUTHORS’ CONTRIBUTIONS

Study conception, data acquisition, synthesis, and first draft: TM. Data acquisition and synthesis: VCM, YW. Resource acquisition: HMB, AF. All authors reviewed and approved the final version for publication.

## COMPETING INTERESTS

The authors have declared that no competing interests exist.

## DATA AVAILABILITY

Data used for all analyses can be found in the Supplementary Files.

## Notes

### Competing Interest Statement

The authors have declared no competing interest.

## REFERENCES

1. Shapiro A, Bradshaw B, Landes S, et al. A novel digital approach to describe real world outcomes among patients with constipation. NPJ Digit Med. 2021;4(1):27.

2. Daly AC, Prendergast ME, Hughes AJ, Burdick JA. Bioprinting for the Biologist. Cell. 2021;184(1):18–32.

3. Zhang Z, Wu HX, Lin WH, et al. EPHA7 mutation as a predictive biomarker for immune checkpoint inhibitors in multiple cancers. BMC Med. 2021;19(1):26.

4. Tschandl P, Rinner C, Apalla Z, et al. Human-computer collaboration for skin cancer recognition. Nat Med. 2020;26(8):1229–1234.

5. Røttingen JA, Farrar J. Targeted health innovation for global health. BMJ. 2019;366:5601

6. Schulman KA, Richman BD. Toward an Effective Innovation Agenda. N Engl J Med. 2019;380(10):900–901.

7. Klonoff DC, King F, Kerr D. New Opportunities for Digital Health to Thrive. J Diabetes Sci Technol. 2019;13(2):159–163.

8. Navarro-Torné A, Hanrahan F, Kerstiëns B, et al. Public Health-Driven Research and Innovation for Next-Generation Influenza Vaccines, European Union. Emerg Infect Dis. 2019;25(x2).

9. Brown EL, Ruggiano N, et al. Smartphone-Based Health Technologies for Dementia Care: Opportunities, Challenges, and Current Practices. J Appl Gerontol. 2019;38(1):73–91.

10. Pham L, Harris T, Varosanec M, Morgan V, Kosa P, Bielekova B. Smartphone-based symbol-digit modalities test reliably captures brain damage in multiple sclerosis. NPJ Digit Med. 2021;4(1):36.

11. Shapiro A, Bradshaw B, Landes S, et al. A novel digital approach to describe real world outcomes among patients with constipation. NPJ Digit Med. 2021;4(1):27.

12. Kvedar JC, Fogel AL. mHealth advances clinical research, bit by bit. Nat Biotechnol. 2017;35(4):337–339.

13. Seto E, Smith D, Jacques M, Morita PP. Opportunities and Challenges of Telehealth in Remote Communities: Case Study of the Yukon Telehealth System. JMIR Med Inform. 2019;7(4):e11353.

14. Dunn P, Hazzard E. Technology approaches to digital health literacy. Int J Cardiol. 2019;293:294–296.

15. Burlone S, Moore L, Johnson W. Overcoming Barriers to Accessing Obstetric Care in Underserved Communities. Obstet Gynecol. 2019;134(2):271–275.

16. Liao Y, Thompson C, Peterson S. The Future of Wearable Technologies and Remote Monitoring in Health Care. Am Soc Clin Oncol Educ Book. 2019;39:115–121.

17. Piao C, Terrault NA, Sarkar S. Telemedicine: An Evolving Field in Hepatology. Hepatol Commun. 2019;3(5):716–721.

18. Fogel AL, Kvedar JC. Artificial intelligence powers digital medicine. NPJ Digit Med. 2018;1:5.

19. Young AT, Fernandez K, Pfau J, et al. Stress testing reveals gaps in clinic readiness of image-based diagnostic artificial intelligence models. NPJ Digit Med. 2021;4(1):10.

20. Randhawa GK, Jackson M. The role of artificial intelligence in learning and professional development for healthcare professionals. Healthc Manage Forum. 2020;33(1):19–24.

21. Wiljer D, Hakim Z. Developing an Artificial Intelligence-Enabled Health Care Practice: Rewiring Health Care Professions for Better Care. J Med Imaging Radiat Sci. 2019 [Epub ahead of print]

22. The Era of Pharmacogenomics. Acta Sci Cancer Biol. 2019;3(7):14–16.

23. Ritchie MD, Moore JH, Kim JH. Translational Bioinformatics: Biobanks in the Precision Medicine Era. Pac Symp Biocomput. 2020;25:743–747.

24. Lienhardt C, Nahid P. Advances in clinical trial design for development of new TB treatments: A call for innovation. PLoS Med. 2019;16(3):e1002769.

25. Lehoux P, Roncarolo F, Silva HP, et al. What Health System Challenges Should Responsible Innovation in Health Address? Insights From an International Scoping Review. Int J Health Policy Manag. 2019;8(2):63–75.

26. Félix IB, Guerreiro MP, Cavaco A, et al. Development of a Complex Intervention to Improve Adherence to Antidiabetic Medication in Older People Using an Anthropomorphic Virtual Assistant Software. Front Pharmacol. 2019;10:680.

27. Shakeri Hossein Abad Z, Kline A, Sultana M, et al. Digital public health surveillance: a systematic scoping review. NPJ Digit Med. 2021;4(1):41..

28. Cornell University, INSEAD, and the World Intellectual Property Organization. Global innovation index 2019: Creating health lives – the future of medical innovation. 12th ed. Ithaca, Fontainebleau, and Geneva, 2019. Available at https://www.wipo.int/edocs/pubdocs/en/wipo_pub_gii_2019.pdf

29. United Nations Conference on Trade and Development (UNCTED). Selected sustainable development trends in the least developed countries – 2019. Geneva, 2019. Available at https://unctad.org/en/PublicationsLibrary/aldc2019d1_en.pdf

30. Shahin MH, Abdel-Rahman S, Hartman D. The Patient-Centered Future of Clinical Pharmacology. Clin Pharmacol Ther. 2019. [Epub ahead of print]

31. Barrenho E, Miraldo M, Smith PC. Does global drug innovation correspond to burden of disease? The neglected diseases in developed and developing countries. Health Econ. 2019;28(1):123–143.

32. Burki T. Developing countries in the digital revolution. Lancet. 2018;391(10119):417.

33. Oloruntoba SO, Muchie M. Innovation, Regional Integration, and Development in Africa: Advances in African Economic, Social and Political Development. Springer, 2019.

34. Mackintosh M, Mugwagwa J, Banda G, et al. Health-industry linkages for local health: reframing policies for African health system strengthening. Health Policy Plan. 2018;33(4):602–610.

35. United Nations Conference on Trade and Development (UNCTED). Technology and innovation report – 2018: Harnessing Frontier Technologies for Sustainable Development. Geneve, 2018. Available at https://unctad.org/en/PublicationsLibrary/tir2018_en.pdf

36. Mugabe JO. World Health Organization. Health innovation systems in developing countries: Strategies for building scientific and technological capacities. World Health Organization, 2005.

37. Simpkin V, Namubiru-Mwaura E, Clarke L, et al. Investing in health R & D: where we are, what limits us, and how to make progress in Africa. BMJ Glob Health. 2019;4(2):e001047.

38. Nabyonga-Orem J, Nabukalu JB, Okuonzi SA. Partnership with private for-profit sector for universal health coverage in sub-Saharan Africa: opportunities and caveats. BMJ Glob Health. 2019 Oct 5;4(9):e001193.

39. Olakunde BO, Adeyinka DA, Ozigbu CE, et al. Revisiting aid dependency for HIV programs in SubSaharan Africa. Public Health. 2019;170:57–60.

40. Taura ND, Bolat E, Madichie NO, editors. Digital Entrepreneurship in Sub-Saharan Africa: Challenges, Opportunities and Prospects. Springer, 2019.

41. World Health Organization (WHO). Global Strategy on Digital Health 2020-2025. Genea, Switzerland; WHO 2020. Available at https://www.who.int/docs/default-source/documents/gs4dhdaa2a9f352b0445bafbc79ca799dce4d.pdf.

42. World Health Organization Regional Office for Africa (WHO-AFRO). Health Technologies and Innovations. Available at https://www.afro.who.int/programmes-clusters/HTI

43. World Bank. World Bank in Ethiopia: Overview. World Bank, 2019. Available at https://www.worldbank.org/en/country/ethiopia/overview

44. J. ohn Snow, Inc. (JSI). Ethiopia Launches Digital Health Innovation and Learning Center. August 6th, 2020 NEWS. https://www.jsi.com/ethiopia-launches-digital-health-innovation-and-learning-center/.

45. U.S Food and Drug Administration (FDA). What is Digital Health? MD, USA; FDA 2020. Available at https://www.fda.gov/medical-devices/digital-health-center-excellence/what-digital-health#:~:text=Digital%20health%20technologies%20use%20computing,applications%20as%20a%20medical%20device.

46. Yigezu A, Alemayehu S, Hamusse SD, Ergeta GT, Hailemariam D, Hailu A. Cost-effectiveness of facility-based, stand-alone and mobile-based voluntary counseling and testing for HIV in Addis Ababa, Ethiopia. Cost Eff Resour Alloc. 2020;18:34.

47. Gebremariam KT, Zelenko O, Hadush Z, Mulugeta A, Gallegos D. Could mobile phone text messages be used for infant feeding education in Ethiopia? A formative qualitative study. Health Informatics J. 2020;26(4):2614–2624.

48. Starr N, Gebeyehu N, Tesfaye A, et al. Value and Feasibility of Telephone Follow-Up in Ethiopian Surgical Patients. Surg Infect (Larchmt). 2020;21(6):533–539.

49. Jadhav A, Weis J. Mobile phone ownership, text messages, and contraceptive use: Is there a digital revolution in family planning?. Contraception. 2020;101(2):97–105.

50. Bradley D, Honeyman C, Patel V, et al. Smartphones can be used for patient follow-up after a surgical mission treating complex head and neck disfigurement in Ethiopia: Results from a prospective pilot study. J Plast Reconstr Aesthet Surg. 2020;S1748–6815(20)30522-2.

51. Tadesse Y, Gelagay AA, Tilahun B, Endehabtu BF, Mekonnen ZA, Gashu KD. Willingness to Use Mobile based e-Partograph and Associated Factors among Care Providers in North Gondar Zone, Northwest Ethiopia. Online J Public Health Inform. 2019;11(2):e10.

52. Kassa ZY, Tenaw Z, Astatkie A, et al. Mobile Phone Based Strategies for Preconception Education in Rural Africa. Ann Glob Health. 2019;85(1):101.

53. Kebede AS, Ajayi IO, Arowojolu AO. Effect of enhanced reminders on postnatal clinic attendance in Addis Ababa, Ethiopia: a cluster randomized controlled trial. Glob Health Action. 2019;12(1):1609297.

54. Thomsen CF, Barrie AMF, Boas IM, et al. Health workers’ experiences with the Safe Delivery App in West Wollega Zone, Ethiopia: a qualitative study. Reprod Health. 2019;16(1):50.

55. Jemere AT, Yeneneh YE, Tilahun B, Fritz F, Alemu S, Kebede M. Access to mobile phone and willingness to receive mHealth services among patients with diabetes in Northwest Ethiopia: a cross-sectional study. BMJ Open. 2019;9(1):e021766.

56. Endehabtu B, Weldeab A, Were M, Lester R, Worku A, Tilahun B. Mobile Phone Access and Willingness Among Mothers to Receive a Text-Based mHealth Intervention to Improve Prenatal Care in Northwest Ethiopia: Cross-Sectional Study. JMIR Pediatr Parent. 2018;1(2):e9.

57. Mengesha W, Steege R, Kea AZ, Theobald S, Datiko DG. Can mHealth improve timeliness and quality of health data collected and used by health extension workers in rural Southern Ethiopia?. J Public Health (Oxf). 2018;40(suppl_2):ii74–ii86.

58. Steege R, Waldman L, Datiko DG, Kea AZ, Taegtmeyer M, Theobald S. ‘The phone is my boss and my helper’ - A gender analysis of an mHealth intervention with Health Extension Workers in Southern Ethiopia. J Public Health (Oxf). 2018;40(suppl_2):ii16–ii31.

59. Martindale S, Mableson HE, Kebede B, et al. A comparison between paper-based and m-Health tools for collating and reporting clinical cases of lymphatic filariasis and podoconiosis in Ethiopia. Mhealth. 2018;4:49.

60. Abate A, Kifle M, Sena Okuboyejo S, Mbarika V. A mobile-based telepathology system for a low resource setting in Ethiopia. Applied Computing and Informatics. 2018;14(2):186–91.

61. Shiferaw S, Workneh A, Yirgu R, Dinant GJ, Spigt M. Designing mHealth for maternity services in primary health facilities in a low-income setting - lessons from a partially successful implementation. BMC Med Inform Decis Mak. 2018;18(1):96.

62. Atnafu A, Otto K, Herbst CH. The role of mHealth intervention on maternal and child health service delivery: findings from a randomized controlled field trial in rural Ethiopia. Mhealth. 2017;3:39.

63. Mableson HE, Martindale S, Stanton MC, Mackenzie C, Kelly-Hope LA. Community-based field implementation scenarios of a short message service reporting tool for lymphatic filariasis case estimates in Africa and Asia. Mhealth. 2017;3:28.

64. Medhanyie AA, Spigt M, Yebyo H, et al. Quality of routine health data collected by health workers using smartphone at primary health care in Ethiopia. Int J Med Inform. 2017;101:9–14.

65. Shiferaw S, Spigt M, Tekie M, Abdullah M, Fantahun M, Dinant GJ. The Effects of a Locally Developed mHealth Intervention on Delivery and Postnatal Care Utilization; A Prospective Controlled Evaluation among Health Centres in Ethiopia. PLoS One. 2016;11(7):e0158600.

66. Lund S, Boas IM, Bedesa T, Fekede W, Nielsen HS S & ørensen BL. Association Between the Safe Delivery App and Quality of Care and Perinatal Survival in Ethiopia: A Randomized Clinical Trial. JAMA Pediatr. 2016;170(8):765–771.

67. Kebede M, Zeleke A, Asemahagn M, Fritz F. Willingness to receive text message medication reminders among patients on antiretroviral treatment in North West Ethiopia: A cross-sectional study. MC Med Inform Decis Mak. 2015;15:65.

68. Medhanyie AA, Little A, Yebyo H, et al. Health workers’ experiences, barriers, preferences and motivating factors in using mHealth forms in Ethiopia. Hum Resour Health. 2015;13(1):2.

69. Desta BF, Mohammed H, Barry D, Frew AH, Hepburn K, Claypoole C. Use of mobile video show for community behavior change on maternal and newborn health in rural Ethiopia. J Midwifery Womens Health. 2014;59 Suppl 1:S65–S72.

70. Little A, Medhanyie A, Yebyo H, Spigt M, Dinant GJ, Blanco R. Meeting community health worker needs for maternal health care service delivery using appropriate mobile technologies in Ethiopia [published correction appears in PLoS One. 2014;9(1).

71. Seboka BT, Yilma TM, Birhanu AY. Factors influencing healthcare providers’ attitude and willingness to use information technology in diabetes management. BMC Med Inform Decis Mak. 2021;21(1):24.

72. Berihun B, Atnafu DD, Sitotaw G. Willingness to Use Electronic Medical Record (EMR) System in Healthcare Facilities of Bahir Dar City, Northwest Ethiopia. Biomed Res Int. 2020;2020:3827328.

73. Ahmed MH, Bogale AD, Tilahun B, et al. Intention to use electronic medical record and its predictors among health care providers at referral hospitals, north-West Ethiopia, 2019: using unified theory of acceptance and use technology 2(UTAUT2) model. BMC Med Inform Decis Mak. 2020;20(1):207.

74. Kebede M, Adeba E, Chego M. Evaluation of quality and use of health management information system in primary health care units of east Wollega zone, Oromia regional state, Ethiopia. BMC Med Inform Decis Mak. 2020;20(1):107.

75. Awol SM, Birhanu AY, Mekonnen ZA, et al. Health Professionals’ Readiness and Its Associated Factors to Implement Electronic Medical Record System in Four Selected Primary Hospitals in Ethiopia. Adv Med Educ Pract. 2020;11:147–154.

76. Zeleke AA, Worku AG, Demissie A, et al. Evaluation of Electronic and Paper-Pen Data Capturing Tools for Data Quality in a Public Health Survey in a Health and Demographic Surveillance Site, Ethiopia: Randomized Controlled Crossover Health Care Information Technology Evaluation. JMIR Mhealth Uhealth. 2019;7(2):e10995.

77. Abiy R, Gashu K, Asemaw T, et al. A Comparison of Electronic Medical Record Data to Paper Records in Antiretroviral Therapy Clinic in Ethiopia: What is affecting the Quality of the Data?. Online J Public Health Inform. 2018;10(2):e212.

78. Bramo SS, Agago TA. Utilization Status of Electronic Information Sources (EIS) for HIV/AIDS Care and Treatment in Specialized Teaching Hospitals of Ethiopia, 2016. Ethiop J Health Sci. 2017;27(5):507–514.

79. Dusabe-Richards JN, Tesfaye HT, Mekonnen J, Kea A, Theobald S, Datiko DG. Women health extension workers: Capacities, opportunities and challenges to use eHealth to strengthen equitable health systems in Southern Ethiopia. Can J Public Health. 2016;107(4-5):e355–e361.

80. Samuel S, Bayissa G, Asaminewu S, Alaro T. Electronic Information Sources Access and Use for Healthcare Services in Governmental and Non-Governmental Hospitals of Western Oromia, Ethiopia: A Cross Sectional Study. Ethiop J Health Sci. 2016;26(4):341–350.

81. Tilahun B, Fritz F. Comprehensive evaluation of electronic medical record system use and user satisfaction at five low-resource setting hospitals in ethiopia. JMIR Med Inform. 2015;3(2):e22.

82. Biruk S, Yilma T, Andualem M, Tilahun B. Health Professionals’ readiness to implement electronic medical record system at three hospitals in Ethiopia: a cross sectional study. BMC Med Inform Decis Mak. 2014;14:115.

83. King JD, Buolamwini J, Cromwell EA, et al. A novel electronic data collection system for large-scale surveys of neglected tropical diseases. PLoS One. 2013;8(9):e74570.

84. Biruk K, Abetu E. Knowledge and Attitude of Health Professionals toward Telemedicine in Resource-Limited Settings: A Cross-Sectional Study in North West Ethiopia. J Healthc Eng. 2018;2018:2389268.

85. Xue Y, Liang H, Mbarika V, Hauser R, Schwager P, Kassa Getahun M. Investigating the resistance to telemedicine in Ethiopia. Int J Med Inform. 2015;84(8):537–547.

86. Delaigue S, Morand JJ, Olson D, Wootton R, Bonnardot L. Teledermatology in Low-Resource Settings: The MSF Experience with a Multilingual Tele-Expertise Platform. Front Public Health. 2014;2:233.

87. Network for Peri-operative Critical care (N4PCc)*. Addressing priorities for surgical research in Africa: implementation of a multicentre cloud-based peri-operative registry in Ethiopia. Anaesthesia. 2021;10.1111/anae.15394.

88. Jede F, Brandt T, Gedefaw M, et al. Home-based HPV self-sampling assisted by a cloud-based electronic data system: Lessons learnt from a pilot community cervical cancer screening campaign in rural Ethiopia. Papillomavirus Res. 2020;9:100198.

89. Quinonez SC, Yeshidinber A, Lourie MA, et al. Introducing medical genetics services in Ethiopia using the MiGene Family History App. Genet Med. 2019;21(2):451–458.

90. Brant AR, Hinkle J, Shi S, et al. Artificial Intelligence in Global Ophthalmology: Using Machine Learning to Improve Cataract Surgery Outcomes at Ethiopian Outreaches. J Cataract Refract Surg. 2020;10.1097/j.jcrs.0000000000000407.

91. Shiferaw KB, Tilahun BC, Endehabtu BF. Healthcare providers’ digital competency: a cross-sectional survey in a low-income country setting. BMC Health Serv Res. 2020;20(1):1021.

92. Kalayou MH, Endehabtu BF, Tilahun B. The Applicability of the Modified Technology Acceptance Model (TAM) on the Sustainable Adoption of eHealth Systems in Resource-Limited Settings. J Multidiscip Healthc. 2020;13:1827–1837.

93. Meyer AJ, Armstrong-Hough M, Babirye D, et al. Implementing mHealth Interventions in a Resource-Constrained Setting: Case Study From Uganda. JMIR Mhealth Uhealth. 2020;8(7):e19552.

94. Mugambe RK, Mselle JS, Ssekamatte T, et al. Impact of mhealth messages and environmental cues on hand hygiene practice among healthcare workers in the greater Kampala metropolitan area, Uganda: study protocol for a cluster randomized trial. BMC Health Serv Res. 2021;21(1):88.

95. Aw M, Ochieng BO, Attambo D, et al. Critical appraisal of a mHealth-assisted community-based cardiovascular disease risk screening program in rural Kenya: an operational research study. Pathog Glob Health. 2020;114(7):379–387.

96. Karlyn A, Odindo S, Onyango R, et al. Testing mHealth solutions at the last mile: insights from a study of technology-assisted community health referrals in rural Kenya. Mhealth. 2020;6:43.

97. Ngongo BP, Ochola P, Ndegwa J, Katuse P. The technological, organizational and environmental determinants of adoption of mobile health applications (m-health) by hospitals in Kenya. PLoS One. 2019;14(12):e0225167.

98. Harrington EK, Drake AL, Matemo D, et al. An mHealth SMS intervention on Postpartum Contraceptive Use Among Women and Couples in Kenya: A Randomized Controlled Trial. Am J Public Health. 2019;109(6):934–941.

99. Klingberg A, Sawe HR, Hammar U, Wallis LA, Hasselberg M. m-Health for Burn Injury Consultations in a Low-Resource Setting: An Acceptability Study Among Health Care Providers. Telemed J E Health. 2020;26(4):395–405.

100. Thomas DS, Daly K, Nyanza EC, Ngallaba SE, Bull S. Health worker acceptability of an mHealth platform to facilitate the prevention of mother-to-child transmission of HIV in Tanzania. Digit Health. 2020;v6:2055207620905409.

101. Lee S, Begley CE, Morgan R, Chan W, Kim SY. Addition of mHealth (mobile health) for family planning support in Kenya: disparities in access to mobile phones and associations with contraceptive knowledge and use. Int Health. 2019;11(6):463–471.

102. Kazi AM, Carmichael JL, Hapanna GW, et al. Assessing Mobile Phone Access and Perceptions for Texting-Based mHealth Interventions Among Expectant Mothers and Child Caregivers in Remote Regions of Northern Kenya: A Survey-Based Descriptive Study. JMIR Public Health Surveill. 2017;3(1):e5.

103. Johnson D, Juras R, Riley P, et al. A randomized controlled trial of the impact of a family planning mHealth service on knowledge and use of contraception. Contraception. 2017;95(1):90–97.

104. Thobias J, Kiwanuka A. Design and implementation of an m-health data model for improving health information access for reproductive and child health services in low resource settings using a participatory action research approach. BMC Med Inform Decis Mak. 2018;18(1):45.

105. Ngowi KM, Lyamuya F, Mmbaga BT, et al. Technical and Psychosocial Challenges of mHealth Usage for Antiretroviral Therapy Adherence Among People Living With HIV in a Resource-Limited Setting: Case Series. JMIR Form Res. 2020;4(6):e14649.

106. Alaiad A, Alsharo M, Alnsour Y. The Determinants of M-Health Adoption in Developing Countries: An Empirical Investigation. Appl Clin Inform. 2019;10(5):820–840.

107. Webster P. Patient data in the cloud. Lancet Digital Health. 2019;1(8): E391–E392.

108. Adane K. The Current Status of Cyber Security in Ethiopia. IUP Journal of Information Technology. 2020 Sep 1;16(3):7–19.

109. Manyazewal T, Woldeamanuel Y, Blumberg HM, Fekadu A, Marconi VC. The fight to end tuberculosis must not be forgotten in the COVID-19 outbreak. Nat Med. 2020;26(6):811–812.

110. Wondimu W, Girma B. Challenges and Silver Linings of COVID-19 in Ethiopia - Short Review. J Multidiscip Healthc. 2020;13:917–922.

